# The Efficacy of Cefazolin and Ertapenem combination treatment for Persistent MSSA Bacteraemia: A ten-year multicentre retrospective cohort study

**DOI:** 10.64898/2026.11.04.26367257

**Authors:** Angus Lane, Karen Hay, Sivasaaini Sivakumaran, Heather Dolby, Stuart Winlaw, Daniel Wise, Visai Muruganandah, Sanjana Heraganahally, Nihal-Indir Lalwani, Matthew Eustace, Fiona Chui, Thomas Kelly, Andrew Redmond, Paul R. Chapman, Alex Chaudhuri, Kanthi Vemuri, Andrew Burke, John McNamara, Bridget E Barber, Robert Horvath

**Affiliations:** Infectious Diseases Department, The Prince Charles Hospital; Australian Collaboration in Endocarditis (ACE), The Prince Charles Hospital; Infectious Diseases Department, The Royal Brisbane and Women’s Hospital; School of Clinical Medicine, The University of Queensland; QIMR Berghofer Medical Research; Herston Infectious Diseases Institute (HeIDi), Metro North Hospital and Health Service; Pathology Queensland

**Keywords:** *Staphylococcus aureus*, bacteraemia, endocarditis, cefazolin, combination drug therapy

## Abstract

**Objectives:** Persistent methicillin-susceptible *Staphylococcus aureus* (MSSA) bacteraemia is associated with significant morbidity and mortality. Cefazolin-ertapenem combination therapy has demonstrated synergistic activity in preclinical models and small case series, but comparative clinical evidence is lacking.

**Methods:** We conducted a multicentre retrospective review of all patients aged ≥18 years with persistent MSSA bacteraemia at two Queensland tertiary hospitals between January 2014 and December 2024. The primary outcome was time to blood culture sterilisation, compared between patients who commenced cefazolin-ertapenem combination therapy prior to first negative blood culture and a matched cohort of non-combination-treated patients. Secondary outcomes included 90-day all-cause mortality, 90-day relapse, development of multi-resistant organisms, *Clostridioides difficile* infection, and adverse drug events.

**Results:** A total of 288 episodes of persistent MSSA bacteraemia in 283 patients were identified. Twenty-six patients received cefazolin-ertapenem combination therapy. Among the 14 patients who commenced combination therapy prior to first negative blood culture, median time to sterilisation was 1 day (IQR 1–2), compared with 2 days (IQR 1–3) in the matched non-combination group. Cox regression in the matched cohort (n=114) demonstrated an increased rate of blood culture clearance with combination therapy (HR 1.8; 95% CI 1.2–2.7; *P* =0.004). In the full cohort, ninety-day mortality (20.0% vs 23.0%, P=0.73) and relapse rates (11.5% vs 8.3%, *P* =0.57) did not differ significantly between groups.

**Conclusions:** Cefazolin-ertapenem combination therapy was associated with accelerated blood culture sterilisation in persistent MSSA bacteraemia. Prospective randomised evaluation is warranted.

## Introduction

*Staphylococcus aureus* bacteraemia (SAB) is a leading cause of infectious mortality worldwide, affecting more than one million patients annually with a 90-day all-cause mortality ranging from 15-30%.[1, 2] Optimal management requires early initiation of effective antibiotic therapy, source control, and infectious diseases consultation.[2] Among patients with methicillin-susceptible *S. aureus* (MSSA) bacteraemia, cefazolin has emerged as the preferred beta-lactam agent, offering comparable efficacy to anti-staphylococcal penicillins with a more favourable tolerability profile, including lower rates of acute kidney injury.[3–5]

Despite best-practice management, approximately 15 - 30% of patients develop persistent bacteraemia, defined as ongoing bacteraemia for ≥72 hours despite appropriate therapies [2, 6] Persistent SAB carries a disproportionate mortality burden, with each additional day of bacteraemia estimated to increase 30-day mortality risk by approximately 16%.[7] There is no established standard of care for refractory MSSA bacteraemia beyond optimised dosing and source control.[2] A specific concern with cefazolin in deep seated infection is the inoculum effect, mediated by expression of the blaZ gene, which can attenuate bactericidal activity at the bacterial burdens encountered in endocarditis, epidural abscess, and persistent bacteraemia.[8, 9]

Combination antibiotic strategies have been explored as a means to accelerate bacteraemia clearance in SAB. Adjunctive rifampicin was associated with faster bacteraemia clearance but conferred no survival benefit in the ARREST trial, and combination cloxacillin-fosfomycin was stopped early for futility in the SAFO trial of MSSA bacteraemia.[10, 11] To address the challenge of persistent MSSA infection, cefazolin-ertapenem combination therapy has been proposed as a salvage strategy. The combination exploits complementary binding affinity for distinct penicillin-binding proteins: cefazolin predominantly targets penicillin binding protein 2 (PBP2) while ertapenem has strong affinity for penicillin binding protein 1 (PBP1), together enhancing bactericidal activity in high-burden infections.[12] In vitro synergy has been demonstrated across multiple assay platforms, and further evidence suggests the combination may enhance host immune clearance through immunomodulatory mechanisms, including stimulation of interleukin-1β release from peripheral blood mononuclear cells.[9, 13–15] Rapid blood culture sterilisation following cefazolin-ertapenem salvage therapy has been reported in five small case series of patients with persistent MSSA bacteraemia, including patients with endocarditis and central nervous system (CNS) metastatic disease.[9, 13, 15–17] Across these case series, a total of 70 patients have received beta-lactam–ertapenem combination therapy for persistent MSSA bacteraemia or endocarditis, including 57 treated specifically with cefazolin-ertapenem; however, no matched comparative analysis has been performed. [9, 13, 15–17] We therefore conducted a multicentre retrospective cohort study of cefazolin-ertapenem as salvage treatment for persistent MSSA bacteraemia across two Queensland tertiary centres over a ten-year period.

## Methods

### Study design and population

This multicentre retrospective observational study was conducted at two tertiary hospitals in Queensland, Australia: The Prince Charles Hospital (TPCH) and the Royal Brisbane and Women’s Hospital (RBWH). We included all patients aged ≥18 years with MSSA bacteraemia of ≥72 hours duration between January 2014 and December 2024. Exclusion criteria were: bacteraemia duration less than 72 hours, polymicrobial bacteraemia, and MRSA as the causative organism. Cases were identified from AUSLAB (PJA Solutions Pty Ltd, Pathology Queensland), which captures all pathology tests ordered from public health facilities in Queensland. Clinical data including comorbidities, antibiotic therapy, and imaging were retrieved from patient records. Ninety-day vital status (alive or deceased) and clinical follow-up were verified through the hospital medical record and Queensland Health’s state-wide clinical information systems, which provide cross-facility admission records and linkage to mortality data for patients managed within the Queensland public health system. Ethics approval was granted by the Metro North Health Human Research Ethics Committee (HREC/2025/MNH/11558).

### Definitions and variables

Persistent MSSA bacteraemia was defined as ≥72 hours of positive cultures.[2] Episodes occurring ≥90 days apart were recorded as separate events. Relapse was defined as a positive MSSA blood culture after 72 hours of negativity with new or worsening symptoms within 90 days.[18, 19] Comorbidity was assessed using the Charlson Comorbidity Index (CCI).[20]

### Statistical analysis

The primary outcome was time to first negative blood culture from the index date. For patients receiving cefazolin-ertapenem “combination therapy”, the index date was the date combination therapy was commenced. For comparator patients (described below), the index date was the corresponding matched day of bacteraemia. Secondary outcomes included 90-day all-cause mortality, 90-day MSSA relapse, development of antimicrobial-resistant organisms, *Clostridioides difficile* infection, and adverse drug events ([ADE], captured across the entire admission). Follow-up was defined as time from the index date until death or 90 days, whichever occurred first.

The primary analysis included patients who commenced combination therapy before the date of their first negative blood culture. Patients in whom clearance coincided with the index date were excluded, as the temporal relationship between treatment commencement and microbiological response could not be reliably established. A sensitivity analysis retaining these patients was also performed.

For each combination-treated patient included in the primary analysis, a matched comparator set was identified comprising all patients who remained bacteraemic on the corresponding bacteremia day and who shared the same age group (<50, 50 - <70, ≥70 years), CCI category (0, 1-4, ≥5), and presence of major metastatic foci (bone/joint, cardiac, or CNS involvement). All eligible comparators were included without a fixed matching ratio; where a comparator was eligible for more than one exposed patient, they were included once only, assigned to the risk set corresponding to the longest duration of bacteraemia. The comparator group comprised patients receiving any regimen other than cefazolin-ertapenem, reflecting real-world practice and heterogeneity. Patients who commenced combination therapy after their first negative blood culture remained eligible for selection as comparators, as eligibility was determined using only information available at the matched timepoint

The effect of combination therapy on time to blood culture clearance was estimated using Cox proportional hazards regression with cluster-robust standard errors to account for the matched structure, with the competing risk of death accounted for in a sensitivity analysis. Summary statistics are presented as mean (SD), median (IQR), or n (%) as appropriate; between-group comparisons used Student’s t-test, Wilcoxon rank-sum, chi-square, or Fisher’s exact test. All analyses were performed in Stata (Version 19.5; StataCorp LLC).[21] A two-sided P<0.05 was considered statistically significant.

## Results

### Study Population

A total of 288 eligible admissions for 283 patients with persistent MSSA bacteraemia were identified (Figure 1). Baseline characteristics for the full cohort by any use of cefazolin-ertapenem combination are shown in Table 1a. The mean (SD) age was 62 (18) years, 87 (31%) were female, and the median (IQR) CCI score was 4 (1–6).

**Figure 1:**
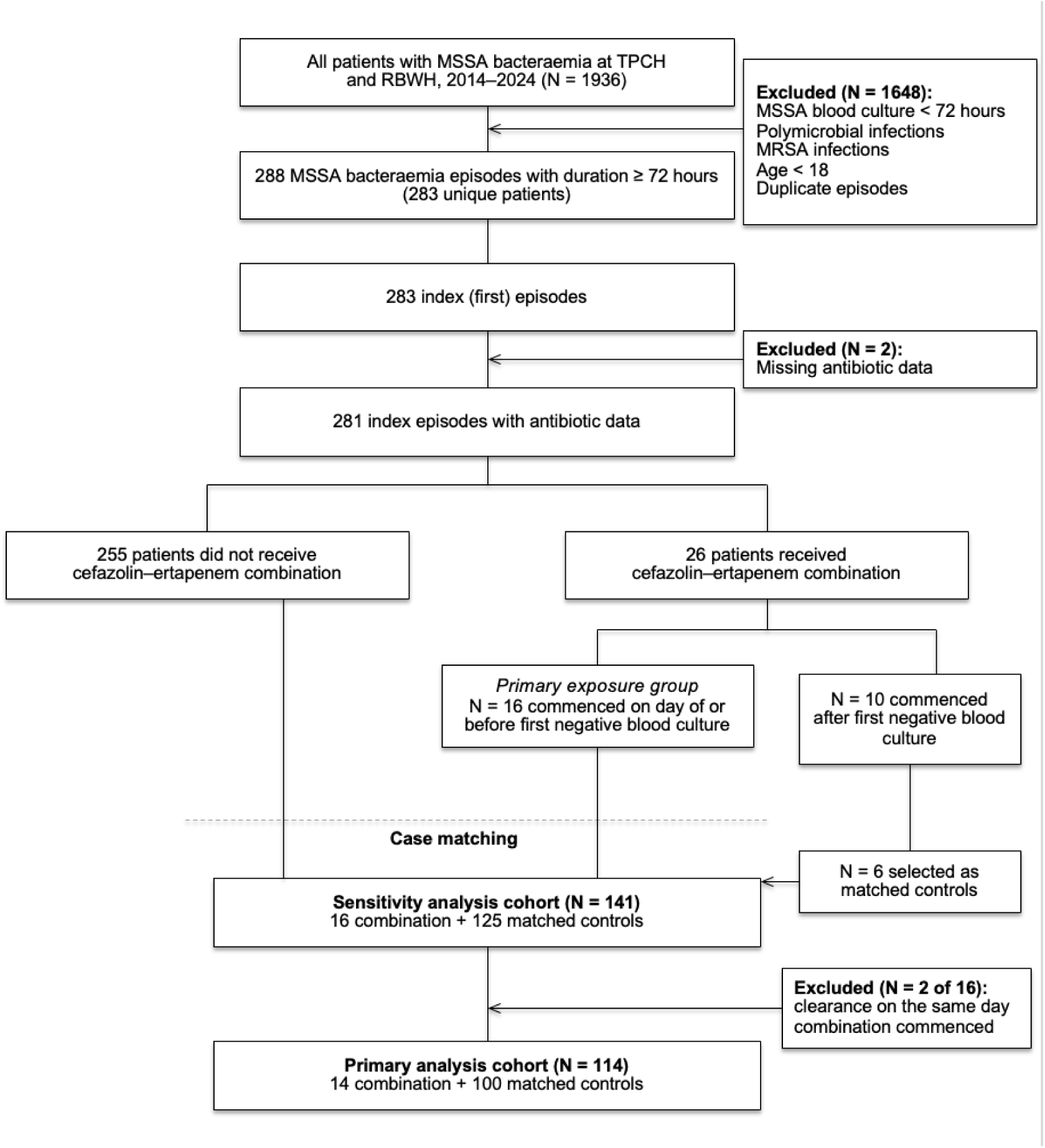
Screening and analysis. MSSA denotes methicillin-susceptible *Staphylococcus aureus*, TPCH The Prince Charles Hospital, RBWH Royal Brisbane and Women’s Hospital. Two patients were excluded from analyses of antibiotic exposure because antibiotic administration dates could not be linked to blood culture dates, leaving 281 index episodes (255 not receiving and 26 receiving cefazolin–ertapenem combination therapy).

**Table 1a.** Baseline Characteristics of the Study Population.

|  | **Other Antibiotic Therapies** | **Cefazolin-Ertapenem Combination therapy** |  |
|---|---|---|---|
| **Variable** | N = 257 | N = 26 | ***P* Value** |
| **Age — years**<sup>\*</sup> | 62 ± 18 | 60 ± 15 | 0.59 |
| **Charlson comorbidity index**<sup>†</sup> | 4 (1–6) | 3 (1–6) | 0.85 |
| **Male sex — no. (%)** | 174 (67.7) | 22 (84.6) | 0.075 |
| **Hospital — no. (%)** |  |  | <0.001 |
| The Prince Charles Hospital | 121 (47.1) | 26 (100.0) | — |
| Royal Brisbane and Women’s Hospital | 136 (52.9) | 0 | — |
| **ICU admission — no. (%)** | 91 (35.8) | 17 (65.4) | 0.003 |
| **Source of bacteraemia — no. (%)** |  |  | 0.82 |
| Skin or soft tissue | 74 (28.8) | 6 (23.1) | — |
| Intravenous drug use | 41 (16.0) | 3 (11.5) | — |
| Intravascular device | 26 (10.1) | 2 (7.7) | — |
| Procedure-related | 16 (6.2) | 1 (3.8) | — |
| Unknown | 100 (38.9) | 14 (53.8) | — |
| **Metastatic focus of infection — no. (%)** |  |  |  |
| Endocarditis |  |  | 0.038 |
| Native valve | 73 (29.6) | 14 (53.8) | — |
| Prosthetic valve | 22 (8.9) | 2 (7.7) | — |
| Bone or joint infection | 99 (39.8) | 17 (65.4) | 0.012 |
| Skin and soft tissue | 65 (26.6) | 7 (26.9) | 0.98 |
| Central nervous system | 47 (19.2) | 9 (36.0) | 0.048 |
| None | 152 (61.5) | 10 (38.5) | — |
| **Diabetes mellitus — no. (%)** |  |  | 0.43 |
| Type 1 | 10 (3.9) | 0 | — |
| Type 2 | 47 (18.3) | 7 (29.2) | — |
| **Chronic kidney disease — no. (%)** | 66 (25.8) | 9 (36.0) | 0.27 |
| **HIV infection — no. (%)**<sup>‡</sup> | 2 (0.9) | 0 | 1.00 |
| **Any immunosuppression — no. (%)** | 114 (44.4) | 13 (50.0) | 0.58 |
| Medication-related | 35 (13.7) | 2 (7.7) | 0.55 |
| Disease-related | 52 (20.3) | 2 (7.7) | 0.19 |
| **Surgery for source control — no. (%)** | 99 (38.5) | 15 (57.7) | 0.058 |
| **Therapeutic drug monitoring performed — no. (%)** | 158 (62.7) | 17 (65.4) | 0.79 |
| **No. of antibiotics received**<sup>†</sup> | 3 (2–4) | 6 (5–7) | <0.001 |

\* Plus–minus values are means ± SD. † Values are medians (interquartile range). All other values are counts (percentage of non-missing).
‡ Calculated among patients with known HIV status (non-combination therapy, N□=□220; combination therapy, N□=□19).
P values are from Student’s t-test (age), Wilcoxon rank-sum test (Charlson comorbidity index; no. of antibiotics received), Pearson’s chi-square test, or Fisher’s exact test, as appropriate. ICU denotes intensive care unit; HIV, human immunodeficiency virus.

**Table 1b.**
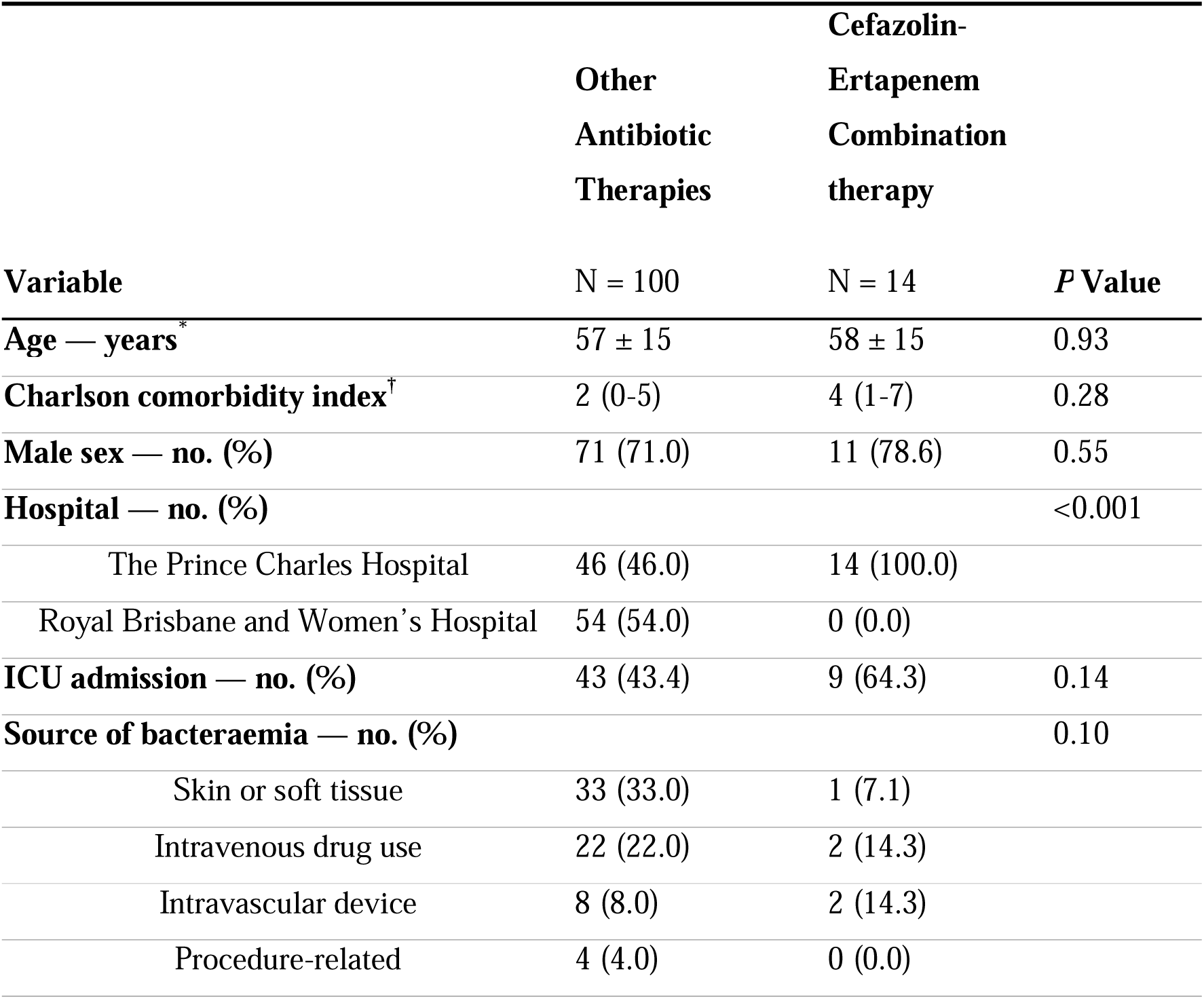
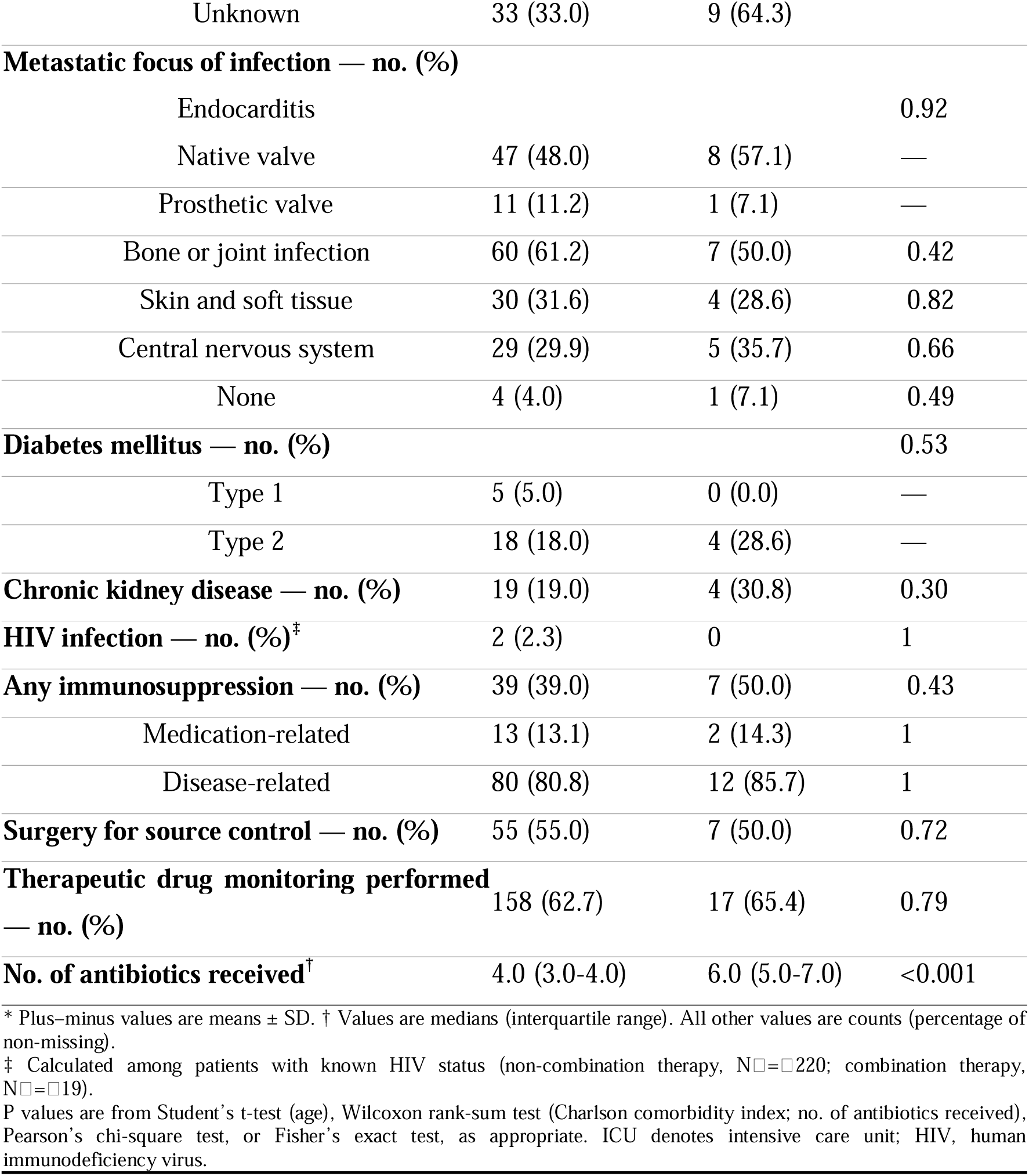
Baseline Characteristics of selected subset by exposure group (N=114).

|  | **Other Antibiotic Therapies** | **Cefazolin-Ertapenem Combination therapy** |  |
|---|---|---|---|
| **Variable** | N = 100 | N = 14 | ***P* Value** |
| **Age — years**<sup>\*</sup> | 57 ± 15 | 58 ± 15 | 0.93 |
| **Charlson comorbidity index**<sup>†</sup> | 2 (0-5) | 4 (1-7) | 0.28 |
| **Male sex — no. (%)** | 71 (71.0) | 11 (78.6) | 0.55 |
| **Hospital — no. (%)** |  |  | <0.001 |
| The Prince Charles Hospital | 46 (46.0) | 14 (100.0) |  |
| Royal Brisbane and Women’s Hospital | 54 (54.0) | 0 (0.0) |  |
| **ICU admission — no. (%)** | 43 (43.4) | 9 (64.3) | 0.14 |
| **Source of bacteraemia — no. (%)** |  |  | 0.10 |
| Skin or soft tissue | 33 (33.0) | 1 (7.1) |  |
| Intravenous drug use | 22 (22.0) | 2 (14.3) |  |
| Intravascular device | 8 (8.0) | 2 (14.3) |  |
| Procedure-related | 4 (4.0) | 0 (0.0) |  |
| Unknown | 33 (33.0) | 9 (64.3) |  |
| **Metastatic focus of infection — no. (%)** |  |  |  |
| Endocarditis |  |  | 0.92 |
| Native valve | 47 (48.0) | 8 (57.1) | — |
| Prosthetic valve | 11 (11.2) | 1 (7.1) | — |
| Bone or joint infection | 60 (61.2) | 7 (50.0) | 0.42 |
| Skin and soft tissue | 30 (31.6) | 4 (28.6) | 0.82 |
| Central nervous system | 29 (29.9) | 5 (35.7) | 0.66 |
| None | 4 (4.0) | 1 (7.1) | 0.49 |
| **Diabetes mellitus — no. (%)** |  |  | 0.53 |
| Type 1 | 5 (5.0) | 0 (0.0) | — |
| Type 2 | 18 (18.0) | 4 (28.6) | — |
| **Chronic kidney disease — no. (%)** | 19 (19.0) | 4 (30.8) | 0.30 |
| **HIV infection — no. (%)**<sup>‡</sup> | 2 (2.3) | 0 | 1 |
| **Any immunosuppression — no. (%)** | 39 (39.0) | 7 (50.0) | 0.43 |
| Medication-related | 13 (13.1) | 2 (14.3) | 1 |
| Disease-related | 80 (80.8) | 12 (85.7) | 1 |
| **Surgery for source control — no. (%)** | 55 (55.0) | 7 (50.0) | 0.72 |
| **Therapeutic drug monitoring performed — no. (%)** | 158 (62.7) | 17 (65.4) | 0.79 |
| **No. of antibiotics received**<sup>†</sup> | 4.0 (3.0-4.0) | 6.0 (5.0-7.0) | <0.001 |

\* Plus–minus values are means ± SD. † Values are medians (interquartile range). All other values are counts (percentage of non-missing).
‡ Calculated among patients with known HIV status (non-combination therapy, N□=□220; combination therapy, N□=□19).
P values are from Student’s t-test (age), Wilcoxon rank-sum test (Charlson comorbidity index; no. of antibiotics received), Pearson’s chi-square test, or Fisher’s exact test, as appropriate. ICU denotes intensive care unit; HIV, human immunodeficiency virus.

Cefazolin-ertapenem combination therapy was administered in 26 patients (9.2%) during their initial episode of bacteremia; one patient received combination therapy during a second episode. Patients with significant metastatic disease and those admitted to the intensive care unit (ICU) were more likely to receive combination therapy.

Baseline characteristics for the selected cohort by use of cefazolin-ertapenem combination therapy prior to first negative blood culture are shown in Table 1b. The selected cohort included in the primary outcome analyses comprised 14 patients who received cefazolin-ertapenem combination therapy prior to negative culture (exposure group) and 100 with matched covariate patterns. The comparator group received a range of antibiotic regimens during their admission. At the matched index date, the most common regimens in the comparator group were flucloxacillin monotherapy (n=42), cefazolin monotherapy (n=5), and flucloxacillin-based combination therapy (n=36), with a small number receiving rifampicin-based combinations (n=2) or other regimens (n=15). Over the full culture-positive period, flucloxacillin was received by 104 (83%) and cefazolin by 32 (26%) of comparator patients, with vancomycin (74%) and gentamicin (33%) predominantly reflecting empiric therapy administered prior to the index timepoint. Further antibiotic regimen information is in Supp Tables 1a and 1b.

Of the 283 patients, 264 (93%) achieved blood culture sterilisation, 15 (5%) died prior to clearance, and 4 were lost to follow-up.

### Cefazolin and Ertapenem Combination Therapy Cohort

Cefazolin and ertapenem combination therapy was first used in 2019, with most cases managed from 2021 onwards (Supp. Table 2). All 26 patients who received combination therapy during their first episode were treated at TPCH. Ten of these patients commenced combination therapy after their first negative blood culture and were therefore not included in the exposure group; as eligibility was assessed at the matched timepoint, these patients remained eligible as comparators, and six were selected as matched controls. Of the remaining 16, two achieved clearance on the date of combination commencement and were similarly excluded from the primary analysis. For the 14 patients included in the primary analysis, the median (IQR) interval from first positive culture to combination commencement was 6 (4.5–9.5) days (range 3–10).

### Primary Outcome

Median (IQR) time to clearance from first positive culture in the overall cohort was 6 (5–9) days (Figure 2). For the primary analysis, a matched comparator group of 100 patients was identified for the 14 evaluable combination-treated patients. Among those who achieved clearance, the median (IQR) time to blood culture sterilisation from the index date was 1 day (1–2; range 1–4) in the combination group and 2 days (1–3; range 1–31) in the comparator group. Cox regression in the matched cohort (n=114) demonstrated an increased rate of clearance with combination therapy (HR 1.8; 95% CI 1.2–2.7; *P*=0.004), with consistent results after accounting for the competing risk of death (Figure 2). In a sensitivity analysis retaining patients with clearance on the index date (n=16 exposed, n=125 comparators), results were consistent (HR 1.45; 95% CI 1.0–2.0; *P* =0.029).

**Figure 2:**
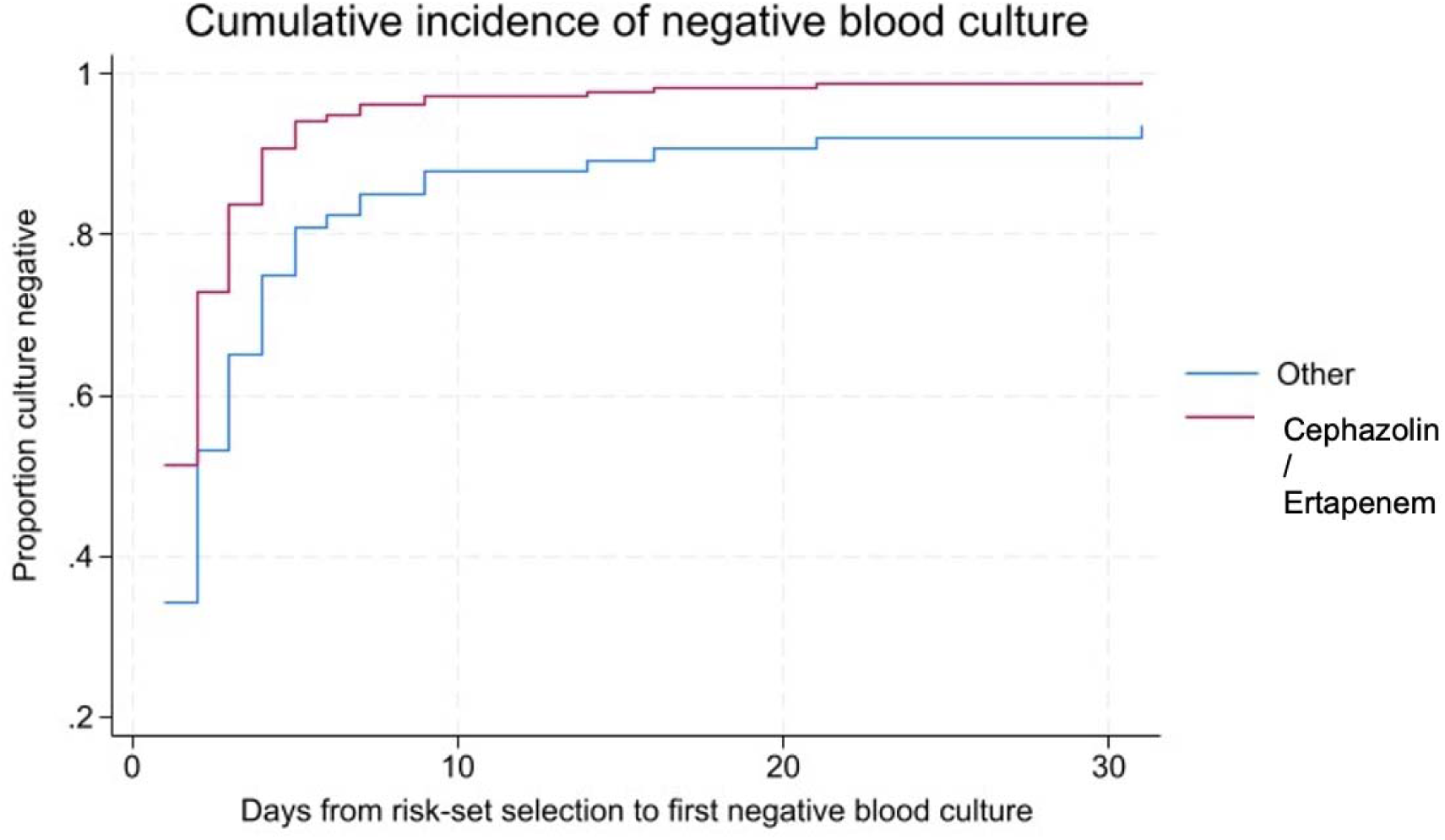
Estimated cumulative incidence of blood culture clearance by group (n=14 treated with ertapenem and cefazolin and n=100 matched on covariate group and time since positive culture). Cumulative incidence functions account for the competing risk of death prior to blood culture clearance. Curves do not reach 1.0, as patients who died before achieving clearance are not recorded as reaching the primary endpoint; the extended plateau in the comparator group reflects a subset of patients with prolonged bacteraemia (range up to 31 days).

### Secondary Outcomes

Ninety-day all-cause mortality (20.0% vs 23.0%; *P* =0.73) and relapse rates (11.5% vs 8.3%; *P* =0.57) did not differ significantly between the combination and comparator groups (Table 2). Adverse drug reactions occurred in 13 (39%) of the 33 patients who received ertapenem across all episodes, most commonly renal (n=6) and hepatic (n=4). However, as events were captured across the admission episode rather than from ertapenem commencement, pre-combination events may have been included, limiting attribution to ertapenem specifically. There was no significant difference in adverse event rates between groups. The median duration of combination therapy was 8 days (IQR 6–15). Antibiotic use during the culture-positive period is detailed in Supp. Table 1.

**Table 2.** Primary and Secondary Outcomes.

| Outcome | Non-combination therapy | Combination therapy | *P* Value |
|---|---|---|---|
| Primary Outcome — Matched Cohort (n□=□114; 14 combination-treated, 100 non-combination-treated)<sup>§</sup> |  |  |  |
| Median days to blood culture clearance from index date<sup>†</sup> | 2 (1–3) | 1 (1–2) | — |
| Hazard ratio for clearance (95% CI)<sup>‡</sup> |  | 1.8 (1.2–2.7) | 0.004 |
| Secondary Outcomes — Matched Cohort |  |  |  |
|  | N□=□100 | N□=□14 |  |
| In-hospital death — no. (%) | 14 (14.0) | 2 (14.3) | 1 |
| 90-day all-cause mortality — no. (%) | 19 (19.0) | 2 (14.3) | 1 |
| 90-day relapse — no. (%) | 7 (7.0) | 1 (7.1) | 1 |
| Adverse drug reaction — no. (%) | 29 (31.2) | 4 (28.6) | 1 |
| New multi-resistant organism — no. (%) | 5 (5) | 1 (7.1) | 0.55 |
| Secondary Outcomes — Full Cohort |  |  |  |
|  | N□=□257 | N□=□26 |  |
| In-hospital death — no. (%) | 50 (19.5) | 4 (15.4) | 0.61 |
| 90-day all-cause mortality — no. (%) | 58 (23.0) | 5 (20.0) | 0.73 |
| 90-day relapse — no. (%) | 21 (8.3) | 3 (11.5) | 0.57 |
| Adverse drug reaction — no. (%) | 75 (30.7) | 10 (38.5) | 0.42 |
| *Clostridioides difficile* infection — no. (%) | 4 (1.6) | 0 (0) | 0.52 |
| New multi-resistant organism — no. (%) | 10 (3.9) | 3 (11.5) | 0.15 |

† Medians (interquartile range). Range: 1–4 days (combination therapy) and 1–31 days (non-combination therapy). Index date is defined as the date of combination therapy commencement (exposed) or the matched equivalent date (controls).
‡ Hazard ratio estimated by Cox proportional-hazards regression with cluster-robust standard errors for matched risk sets, adjusting for age band, Charlson comorbidity category, and major metastatic focus. In a sensitivity analysis including day-0 clearance cases and applying a 0.5-day convention (n□=□141; 16 combination-treated, 125 non-combination-treated), the hazard ratio was 1.45 (95% CI 1.0–2.0; P□=□0.029).
§ Patients commencing combination therapy on the date of first negative blood culture (day-0 clearance, n□=□2) were excluded from the primary analysis. All eligible controls were included without a fixed matching ratio; each control contributed to the longest risk set for which they were eligible. CI denotes confidence interval.
P values for secondary outcomes are from Fisher's exact test or Pearson's chi-square test, as appropriate.

## Discussion

In this retrospective cohort study, cefazolin-ertapenem combination therapy was associated with accelerated blood culture sterilisation in patients with persistent MSSA bacteraemia, with a median time to clearance of one day from the index date compared with two days in matched comparator patients (HR 1.8; 95% CI 1.2–2.7; P=0.004). The rapidity of sterilisation is consistent with prior case series, in which clearance within 24 hours was observed in the majority of evaluable patients.[9, 13, 15] Although each additional day of bacteraemia is estimated to increase 30-day mortality risk by approximately 16%, randomised trials of strategies that accelerate clearance have not consistently translated this association into a mortality benefit; whether cefazolin-ertapenem combination therapy can do so remains an open question.[6, 22] Randomised trials of combination antibiotic therapy in SAB have demonstrated shorter bacteraemia duration without mortality benefit, a pattern observed with vancomycin-flucloxacillin in MRSA bacteraemia (CAMERA2) and cloxacillin-fosfomycin in MSSA bacteraemia (SAFO), and whether the association seen here reflects a true causal pathway or residual confounding by indication cannot be established from retrospective data.[10, 23]

No significant difference in 90-day mortality was observed (20.0% vs 23.0%), though the study was not powered to detect such a difference. This is reassuring given that combination therapy was preferentially used in the most clinically complex patients, those with significant metastatic disease, ICU-level illness, and greater comorbidity burden, in whom worse outcomes might *a priori* be expected, and further suggests the observed effect on clearance may be conservative. Adverse drug reaction rates were numerically higher in the combination group, though attribution in the retrospective setting is inherently limited given the substantial overlap between manifestations of systemic sepsis and antibiotic toxicity, and the markedly greater number of concurrent antimicrobial agents received by combination-treated patients (median 6 vs 3, P<0.001).[24, 25]

The median duration of combination therapy was 8 days (IQR 6–15), longer than the median of 5 days reported in a recent cohort employing a standardised culture-guided approach in which ertapenem was discontinued once blood cultures had been negative for 48 hours.[17] Shorter, protocol-driven courses may be sufficient to achieve sterilisation while reducing the risk of resistant organism selection, a concern supported by the numerically higher rate of multi-resistant organism acquisition in combination-treated patients (11.5% vs 3.9%, P=0.15), a signal that warrants prospective evaluation.

This study has several important limitations. Treatment allocation was not randomised, and confounding by indication is likely given the preferential use of combination therapy in more refractory and complex cases. While the matched cohort design partially accounted for confounding in the primary analysis, no adjustment for confounders was made for secondary outcomes including 90-day mortality; these results should therefore be interpreted with this caveat in mind. The combination-treated cohort was managed predominantly from 2021 onwards, whereas comparators were drawn from the full ten-year study period; improvements in standard of care over this interval may have differentially influenced outcomes. All 26 combination-treated patients during their first episode were managed at a single centre, limiting generalisability. The small exposed group constrains statistical power, and heterogeneity in dosing, timing, and source control strategies may have further confounded results. This underscores a fundamental limitation of retrospective salvage therapy evaluations: without randomisation, spontaneous bacteraemia resolution cannot be excluded as an alternative explanation for the observed culture clearance. Sex was not analysed as independent predictor of outcome; this represents a limitation to the generalisability of findings given the observed imbalance in sex distribution between groups.

This study has several notable strengths. The cohort comprises 288 episodes of persistent MSSA bacteraemia accrued across two Queensland tertiary hospitals over a ten-year study period, representing one of the largest reported cohorts of this syndrome. The matched cohort design, in which comparators were required to remain bacteraemic on the corresponding matched bacteraemia day and share key prognostic characteristics including age group, comorbidity burden, and presence of major metastatic foci, substantially mitigates confounding by illness severity. The use of all eligible comparators without a fixed matching ratio, and the application of cluster-robust standard errors in the Cox regression model, further strengthens the validity of the primary analysis. Together, these methodological features represent a meaningful advance over prior uncontrolled case series describing salvage combination therapy for persistent MSSA bacteraemia.

## Conclusion

Cefazolin-ertapenem combination therapy was associated with accelerated blood culture sterilisation in persistent MSSA bacteraemia, representing the first matched comparative evidence for this strategy. The present findings add to a growing body of observational evidence supporting this combination, but randomised evaluation is needed to establish whether accelerated clearance translates into improvements in mortality and relapse. The ongoing SNAP trial, which includes a dedicated cefazolin-ertapenem arm (NCT04886284), will be particularly informative in this regard. Future research should additionally evaluate optimal combination duration and patient-level predictors of treatment response.

## Data Availability

All data produced in the present study are available upon reasonable request to the authors

## Acknowledgement of Funding

Nil funding was received for this project.

## Author Contributions

Author contributions are reported in accordance with the CRediT (Contributor Roles Taxonomy) framework. AL: Conceptualisation, methodology, data collection, formal analysis, writing, original draft, writing, review and editing, project administration. SS, HD, DW, VM, SH, NIL, SW, ME, FC: Data curation, writing, review and editing. KH: Formal analysis, methodology, writing, review and editing. BB, AR, PC, AC, KV, AB, JNM: Conceptualisation, methodology, writing, review and editing. RH: Conceptualisation, methodology, supervision, writing, review and editing. All authors reviewed and approved the final manuscript prior to submission.

## AI Use statement

Artificial intelligence (AI) writing assistance tools, specifically Claude (Anthropic), were used during the preparation of this manuscript to support proof-reading and editing of the manuscript. All AI-associated content was critically reviewed, verified against source data and primary literature, and revised by the authors. AI tools were not used for data collection, statistical analysis, or interpretation of results. The authors take full responsibility for the integrity and accuracy of the content of this manuscript.

## Supplementary Materials

### Definitions

With regard to adverse drug reactions, acute kidney injury (AKI) was diagnosed in line with KDIGO creatinine criteria for diagnosing AKI: diagnosed when either of the following serum creatinine changes occur: absolute rise ≥0.3 mg/dL (≥26.5 μmol/L) within 48 hours or, relative rise to ≥1.5× baseline within 7 days.[27] Haematologic toxicity was defined as new-onset neutropenia (absolute neutrophil count <1.0 × 10C/L) or thrombocytopenia (platelet count <100 × 10C/L) arising during the treatment period. Muscle toxicity was defined as a creatine kinase (CK) elevation greater than the upper limit of normal, or clinically documented myopathy, where daptomycin formed part of the treatment regimen. Central nervous system toxicity was subcategorised as drug-attributable delirium, defined by clinical documentation in the medical record in the absence of an infective or metabolic aetiology, or new-onset seizure occurring during the treatment period without a prior seizure history or alternative precipitant. Gastrointestinal toxicity included new diarrhoea attributed to antibiotic use, as well as *Clostridioides difficile* infection confirmed by toxin enzyme immunoassay or nucleic acid amplification testing within 90 days of antibiotic commencement. Hypersensitivity reactions were defined as any immune-mediated response to an antimicrobial agent, including urticaria, angioedema, bronchospasm, or anaphylaxis, as documented by the treating clinician. Cutaneous reactions encompassed drug-attributable rash of any morphology not meeting criteria for hypersensitivity.

Immunosuppression was defined as the presence of any of the following: chronic kidney disease, Human immunodeficiency virus (HIV) infection, diabetes mellitus (type 1 or type 2), medication-related immunosuppression (see below) or disease-related immunosuppression (see below). Medication-related immunosuppression was defined as current or recent use of any of the following agents: tumour necrosis factor-alpha (TNF-α) antagonists; interleukin-6 (IL-6) antagonists (including tocilizumab); leflunomide; methotrexate; kinase inhibitors (including imatinib and ibrutinib); calcineurin inhibitors (ciclosporin and tacrolimus); mammalian target of rapamycin (mTOR) inhibitors (sirolimus and everolimus); azathioprine; mycophenolate; cytotoxic chemotherapy; corticosteroids at a dose of ≥20 mg prednisolone equivalent per day for ≥3 weeks; anti-CD52 monoclonal antibody therapy (alemtuzumab); anti-CD20 monoclonal antibody therapy (rituximab); proteasome inhibitors (bortezomib and carfilzomib); or thalidomide analogues (lenalidomide, thalidomide, and pomalidomide). Disease-related immunosuppression was defined as the presence of any of the following: an established autoimmune or inflammatory condition requiring ongoing medical management; active solid organ malignancy, excluding non-melanoma skin cancer; haematological malignancy, regardless of treatment status; or a primary immunodeficiency disorder of systemic significance.

Metastatic foci of infection were identified from clinical documentation, imaging, and microbiological records, and categorised by anatomical site as follows. Endocarditis was classified as native valve endocarditis or prosthetic valve endocarditis, based on modified Duke criteria. Cases in which endocarditis status could not be determined from available records were classified as unknown. Bone and joint infection were subcategorised as native joint infection, prosthetic joint infection, infection involving multiple joints, osteomyelitis without joint involvement, or combined osteomyelitis with native joint infection. Cases in which bone or joint involvement could not be determined were classified as unknown. Skin and soft tissue infection was recorded as present or absent, with cases in which this could not be determined classified as unknown. CNS metastatic foci were defined as microbiologically or radiologically confirmed infection at any intracranial or intraspinal site, and subcategorised as brain abscess, cerebral emboli, epidural abscess, or combined cerebral and epidural disease. Cases in which CNS involvement could not be determined were classified as unknown.

**Supplementary Table 1a.** Antibiotic Use at Index Date in Matched Cohort (N=114).

|  | **Matched cohort** |
|---|---|
| **Regimen at index date** | N = 114 |
| Flucloxacillin monotherapy — no. (%) | 42 (36.8) |
| Cefazolin monotherapy | 5 (4.4) |
| **Rifampicin-based regimens** | 2 (1.8) |
| Rifampicin | 1 (0.9) |
| Rifampicin + other* | 1 (0.9) |
| **Cefazolin–ertapenem combination** | 14 (12.3) |
| Cefazolin + ertapenem | 13 (11.4) |
| Cefazolin + ertapenem + vancomycin | 1 (0.9) |
| **Flucloxacillin-based combinations** | 36 (31.6) |
| Flucloxacillin + vancomycin | 14 (12.3) |
| Flucloxacillin + benzylpenicillin | 2 (1.8) |
| Flucloxacillin + ceftriaxone | 2 (1.8) |
| Flucloxacillin + gentamicin | 2 (1.8) |
| Flucloxacillin + piperacillin–tazobactam | 2 (1.8) |
| Flucloxacillin + vancomycin + ceftriaxone | 2 (1.8) |
| Flucloxacillin + vancomycin + gentamicin | 2 (1.8) |
| Flucloxacillin + vancomycin + piperacillin–tazobactam | 2 (1.8) |
| Flucloxacillin + ertapenem + vancomycin | 2 (1.8) |
| Flucloxacillin + rifampicin | 1 (0.9) |
| Flucloxacillin + other* | 1 (0.9) |
| Flucloxacillin + benzylpenicillin + gentamicin + other* | 1 (0.9) |
| Flucloxacillin + vancomycin + ceftriaxone + metronidazole | 1 (0.9) |
| Flucloxacillin + vancomycin + piperacillin–tazobactam + meropenem + gentamicin | 1 (0.9) |
| Cefazolin + flucloxacillin + vancomycin | 1 (0.9) |
| **Other regimens** | 15 (13.2) |
| Cefazolin + vancomycin | 3 (2.6) |
| Benzylpenicillin | 2 (1.8) |
| Vancomycin | 2 (1.8) |
| Vancomycin + piperacillin–tazobactam | 2 (1.8) |
| Lincomycin | 1 (0.9) |
| Vancomycin + gentamicin + ceftriaxone | 1 (0.9) |
| Vancomycin + meropenem | 1 (0.9) |
| Vancomycin + meropenem + other* | 1 (0.9) |
| Vancomycin + piperacillin–tazobactam + cefepime | 1 (0.9) |
| Vancomycin + lincomycin + meropenem | 1 (0.9) |

Values are counts (percentage) of patients in the matched primary analysis cohort (14 combination-treated, 100 matched comparators); each patient is counted once, under the regimen received on the index date.
\* Other denotes an antibiotic not separately listed.

**Supplementary Table 1b.** Antibiotic Use during the Culture-Positive Period, by Exposure Group.

| **Antibiotic** | **Other Antibiotic Regimens**<br>N = 125 | **Cefazolin-Ertapenem Combination**<br>N = 16 |
|---|---|---|
| Flucloxacillin — no. (%) | 104 (83) | 14 (88) |
| Vancomycin | 92 (74) | 14 (88) |
| Gentamicin | 41 (33) | 7 (44) |
| Piperacillin-tazobactam | 35 (28) | 7 (44) |
| Cefazolin | 32 (26) | 16 (100) |
| Ceftriaxone | 27 (22) | 7 (44) |
| Benzylpenicillin | 9 (7) | 2 (13) |
| Rifampicin | 8 (6) | 0 |
| Metronidazole | 8 (6) | 2 (13) |
| Meropenem | 6 (5) | 1 (6) |
| Ertapenem | 3 (2) | 16 (100) |
| Lincomycin | 3 (2) | 2 (13) |
| Clindamycin | 2 (2) | 1 (6) |
| Linezolid | 2 (2) | 0 |
| Amoxicillin-clavulanic acid | 1 (1) | 0 |
| Cefepime | 1 (1) | 0 |
| Ceftaroline | 1 (1) | 0 |
| Daptomycin | 1 (1) | 0 |
| Ciprofloxacin | 0 | 1 (6) |
| Other | 15 (12) | 3 (19) |

Values are numbers (percentage of group). The matched cohort comprises 16 patients who commenced cefazolin-ertapenem combination therapy prior to first negative blood culture and 125 matched controls (sensitivity-analysis cohort, n = 141). Data reflect all antibiotics administered during the index admission, including empirical agents (e.g., a single dose of gentamicin or vancomycin pending susceptibility results) and directed therapy. One patient may have received more than one antibiotic; percentages do not sum to 100. “Other” Antibiotics include.

**Supplementary Table 2.** Distribution of Cefazolin–Ertapenem Combination Therapy, by Year and Commencement Time.

| **Year** | **Combination therapy not received** | **Cefazolin–ertapenem combination** |  | **First-episode total, n (%)** | **All SAB episodes, n (combination, n [%])** |
|---|---|---|---|---|---|
|  | “all comers”, N = 255 | Before or on day of first negative culture, N = 16 | After first negative culture, N = 10 | N = 281 |  |
| 2014 | 25 (9.8%) | 0 (0.0%) | 0 (0.0%) | 25 (8.9%) | 25 (0) |
| 2015 | 21 (8.2%) | 0 (0.0%) | 0 (0.0%) | 21 (7.5%) | 21 (0) |
| 2016 | 25 (9.8%) | 0 (0.0%) | 0 (0.0%) | 25 (8.9%) | 25 (0) |
| 2017 | 22 (8.6%) | 0 (0.0%) | 0 (0.0%) | 22 (7.8%) | 22 (0) |
| 2018 | 30 (11.8%) | 0 (0.0%) | 0 (0.0%) | 30 (10.7%) | 30 (0) |
| 2019 | 29 (11.4%) | 1 (6.2%) | 0 (0.0%) | 30 (10.7%) | 30 (0) |
| 2020 | 23 (9.0%) | 0 (0.0%) | 2 (20.0%) | 25 (8.9%) | 26 (1 (4%)) |
| 2021 | 25 (9.8%) | 4 (25.0%) | 1 (10.0%) | 30 (10.7%) | 32 (5 (16%)) |
| 2022 | 19 (7.5%) | 5 (31.2%) | 1 (10.0%) | 25 (8.9%) | 25 (6 (24%)) |
| 2023 | 20 (7.8%) | 5 (31.2%) | 1 (10.0%) | 26 (9.3%) | 27 (7 (26%)) |
| 2024 | 16 (6.3%) | 1 (6.2%) | 5 (50.0%) | 22 (7.8%) | 23 (6 (26%)) |
| Total | 255 (90.7%) | 16 (5.7%) | 10 (3.6%) | 281 (100.0%) | 286 (27 (9%)) |

Values are numbers (percentage of column N). Combination therapy not received includes all consecutive patients with methicillin-susceptible Staphylococcus aureus bacteraemia who did not receive cefazolin–ertapenem at any point during their admission (all comers, N = 255). Before first negative culture denotes the primary exposure group, who commenced combination therapy prior to blood culture clearance (N = 16). After first negative culture denotes patients who commenced combination after the first negative blood culture (N = 10). The first-episode total reflects each year's contribution to the 281-patient index-episode cohort. All SAB episodes includes both index and repeat bacteraemia admissions across the study period; the number of episodes in which cefazolin–ertapenem was used is shown in parentheses.

**Supp. Figure 1:**
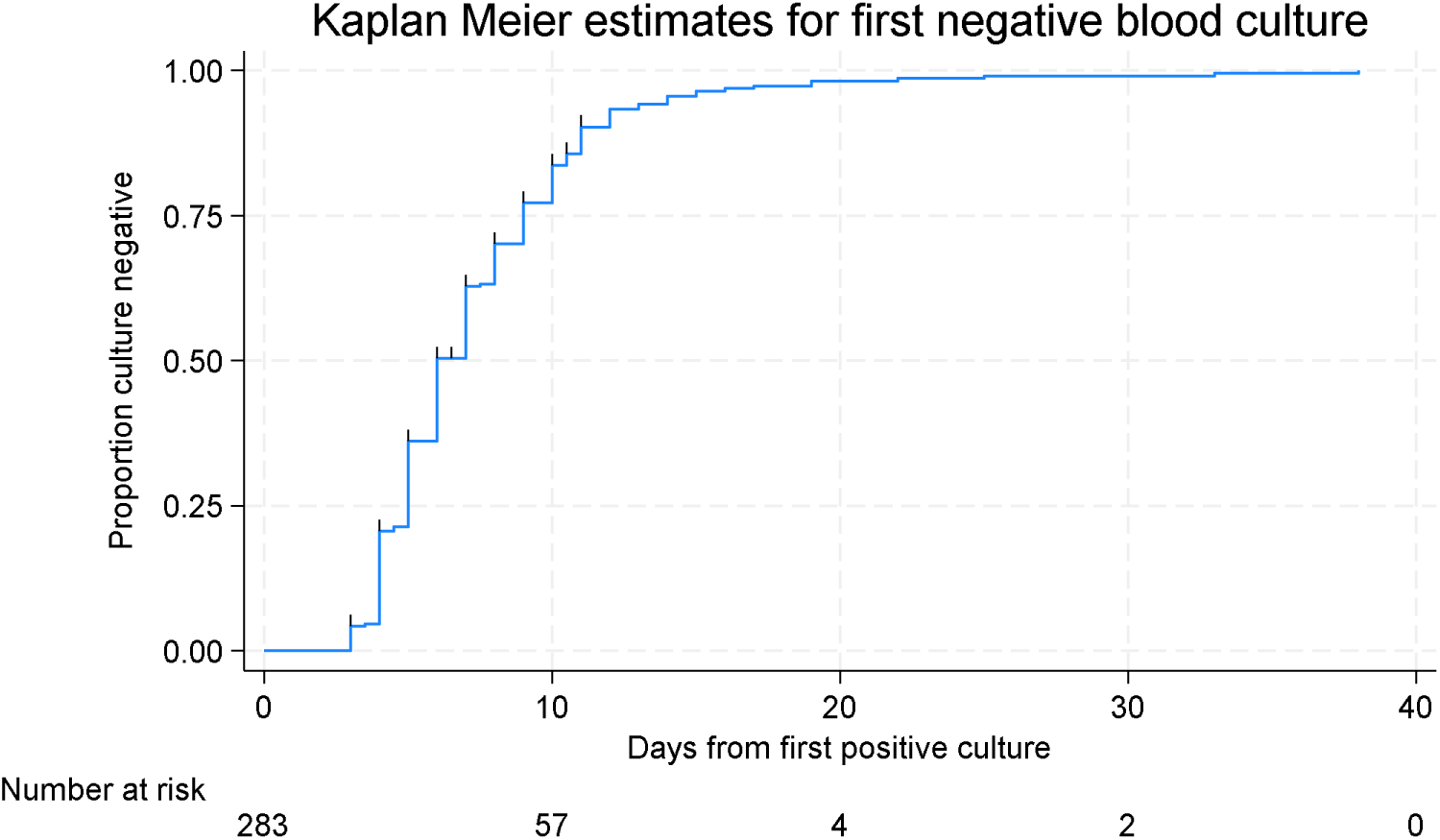
Kaplan Meier estimate for time to first negative culture in full sample (ticks indicate censoring)

**Supplementary Table 3.** Exploratory Subgroup Analyses.

|  | **Exposed group** | **Comparison group** | **p-value** |
|---|---|---|---|
|  | Combination, N = 15 | Non-combination, N= 94 |  |
| Dead at 90 days, n (%) | 2 (13.3) | 25 (25.8) | 0.35 |
| Alive at 90 days, n | 13 (86.7) | 69 (73.4) | — |
|  | Combination, N = 10 | Non-combination, N = 148 |  |
| Dead at 90 days, n (%) | 3 (30.0) | 28 (18.9) | 0.413 |
| Alive at 90 days, n | 7 (70.0) | 120 (81.1) | — |

All analyses are exploratory and hypothesis-generating only. p-values are two-sided from Fisher's exact test;. Endocarditis subgroup includes patients with native or prosthetic valve endocarditis (n = 111); the non-endocarditis subgroup (n = 162) is provided for context (n=10 unknown excluded). Exposed group for the endocarditis and non-endocarditis panels denotes patients who received cefazolin–ertapenem combination therapy; comparison group denotes those who did not.

